# A Novel Phenotype-based Approach for Prioritizing Candidate Genetic Variants for Autism Spectrum Disorder

**DOI:** 10.64898/2026.06.29.26356904

**Authors:** Noam Levi, Mor Dekel, Michal Ilan, Dikla Zigdon, Analya Michaelovsky, Tamar Kolodny, Gal Meiri, Idan Menashe

**Affiliations:** Department of Epidemiology, Biostatistics and Community Health Sciences, Faculty of Health Sciences, Ben-Gurion University of the Negev, Beer Sheva, Israel; Azrieli National Centre for Autism and Neurodevelopment Research, Ben-Gurion University of the Negev, Beer Sheva, Israel; Preschool Psychiatric Unit, Soroka University Medical Center, Beer Sheva, Israel; Child Development Center, Soroka University Medical Center, Beer Sheva, Israel; Psychology Department, Ben-Gurion University of the Negev, Beer Sheva, Israel

## Abstract

**Purpose:** Autism spectrum disorder (ASD) is genetically and phenotypically heterogeneous condition, complicating identification of causal variants. Current diagnostic approaches, have limited diagnostic yield, underscoring the need for new strategies.

**Methods:** We developed a phenotype-driven framework to prioritize ASD-associated genetic variants using comprehensive phenotypic and exome sequencing (ES) data from 125 children with ASD. We used the Human Phenotype Ontology (HPO) nomenclature to prioritize candidate variants in each child based on the similarity between its observed phenotypes and variant-specific expected phenotypes.

**Results:** We identified 228 HPO terms grouped into 41 phenotype categories. ASD-associated genes, according to HPO and SFARI Gene databases, were significantly enriched with these phenotypes compared to non-ASD genes (mean 16.1±5.7 vs. 6.5±5.4; p=1.1^e-231^; HPO, and 16.0±6.7 vs. 7.3±6.0; p=2.1^e-57^; SFARI) supporting the relevance of this phenotype battery to ASD genetics. In 36 genetically resolved participants, the phenotype-similarity approach ranked 58% of causal variants first and 89% within the top three. In the 89 unresolved cases, it highlighted six novel clinically relevant variants, thus increasing the diagnostic yield by 45%.

**Conclusions:** Our novel ASD phenotype battery facilitates prioritization of clinically relevant ASD variants and hence may enhance diagnostic yield, gene discovery, and phenotype-guided precision medicine of ASD.

## Introduction

Autism spectrum disorder (ASD) is a heterogeneous neurodevelopmental condition defined by persistent deficits in social communication and the presence of restricted, repetitive behaviors^1^. Beyond these core domains, most individuals with ASD exhibit a wide range of co-occurring features, including cognitive, behavioral, and medical comorbidities, which complicate both diagnosis and research. This substantial phenotypic variability is widely thought to reflect the underlying etiologic heterogeneity of ASD^2–4^.

ASD is among the most heritable neurodevelopmental disorders^5^, underscoring a major role for genetic factors in its etiology. Early genetic studies identified only a limited number of associated variants; however, the advent of next-generation sequencing (NGS), particularly whole-exome sequencing (ES), has transformed the field by enabling the discovery of numerous rare coding variants and risk genes implicated in ASD^6,7^. Despite these advances, clinically relevant are currently identified in only a subset of individuals, typically 3% to 28%, leaving a substantial proportion of the genetic architecture unresolved^8–10^.

Integrative approaches that systematically link genotype to phenotype offer a promising avenue to improve variant interpretation and diagnostic yield^11^. Accordingly, several computational tools and databases have been developed to support phenotype-driven gene prioritization. For example, Exomiser leverages structured phenotypic data from the HPO to enable automated, high-throughput gene ranking^12,13^, whereas VarElect utilizes the GeneCards knowledgebase to evaluate the relevance of genes and variants to specific phenotypes and diseases^14,15^.

However, the performance of phenotype-driven prioritization approaches critically depends on the accuracy, depth, and relevance of the phenotypic input. This limitation is particularly pronounced in ASD, where clinical heterogeneity and variable phenotypic documentation may reduce the sensitivity of existing tools. To address this gap, we developed a comprehensive, ASD-specific standardized phenotype battery designed to systematically capture clinically relevant features and enhance the prioritization of disease-causing variants, thereby facilitating the identification of clinically actionable genetic findings in individuals with ASD.

## Methods

### Study sample

The study sample was drawn from the database of the Azrieli National Centre for Autism and Neurodevelopment Research (ANCAN)^16^, which includes data on 2,805 children diagnosed with ASD and their families (data freeze: March 2025; Table 1). This database comprises a comprehensive set of sociodemographic and clinical information, including behavioral evaluations, cognitive assessments, developmental milestones, adaptive functioning measures, sensory profiles, and sleep characteristics. Approximately one-third of the children in ANCAN are enrolled in its genetic study, which involves ES of family trios (children with ASD and their parents)^17^. For this study, we randomly selected 100 children with ASD treated at Soroka University Medical Center (SUMC)^18^ for whom ES data was available. Of these, 11 children had known clinically relevant variants and hence considered genetically resolved cases (genetic diagnostic yield of 11%). Another 25 genetically resolved cases from SUMC were added to this sample to enhance statistical power of analyses. Consequently, the study sample comprised of 125 children for whom 36 were genetically resolved.

**Table 1.**
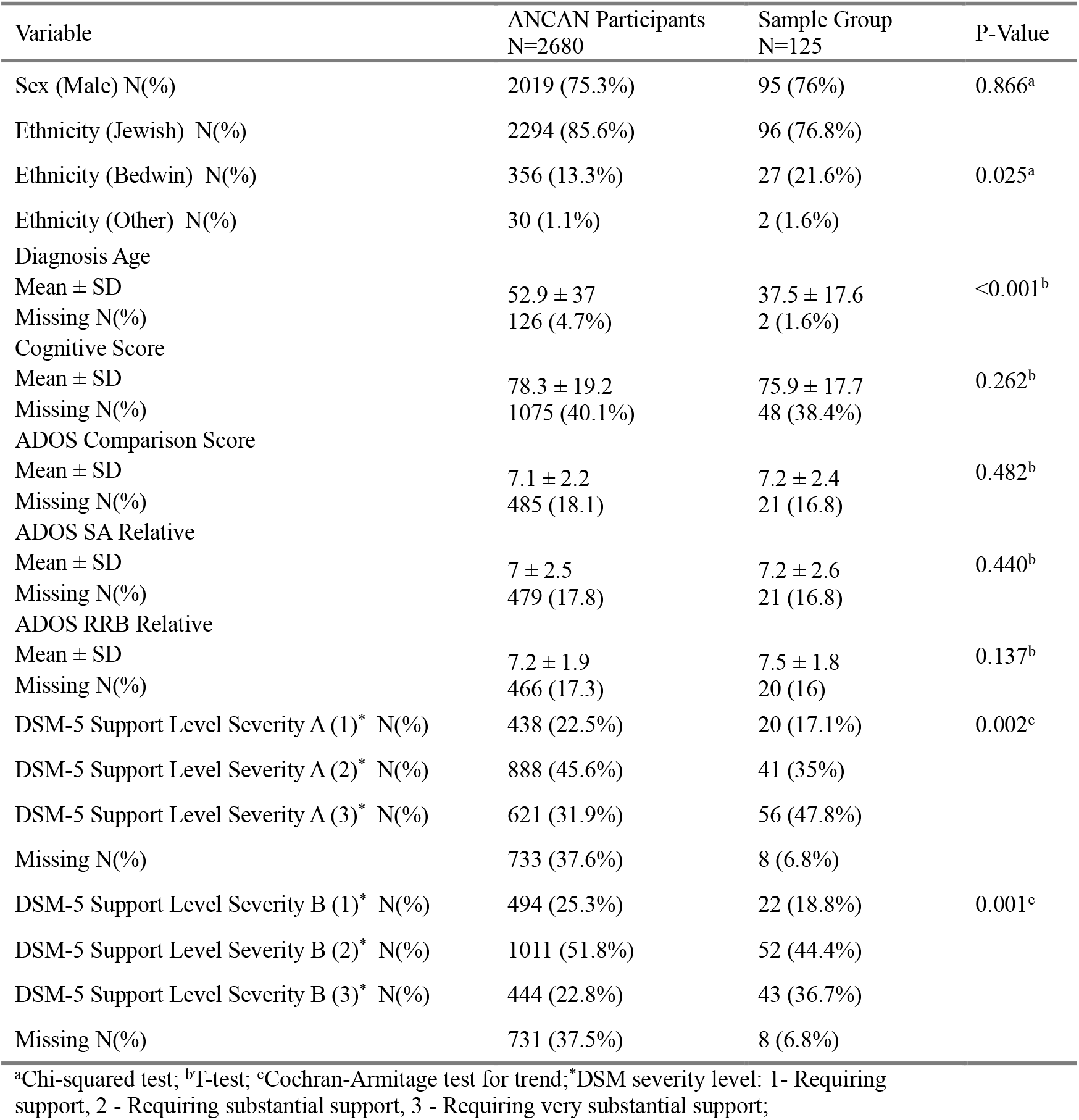
Comparison of Sociodemographic and Clinical Characteristics Among the Sample Group and the rest of ANCAN participants.

| Variable | ANCAN Participants<br>N=2680 | Sample Group<br>N=125 | P-Value |
| --- | --- | --- | --- |
| Sex (Male) N(%) | 2019 (75.3%) | 95 (76%) | 0.866<sup>a</sup> |
| Ethnicity (Jewish) N(%) | 2294 (85.6%) | 96 (76.8%) |  |
| Ethnicity (Bedwin) N(%) | 356 (13.3%) | 27 (21.6%) | 0.025<sup>a</sup> |
| Ethnicity (Other) N(%) | 30 (1.1%) | 2 (1.6%) |  |
| Diagnosis Age |  |  |  |
| Mean ± SD | 52.9 ± 37 | 37.5 ± 17.6 | <0.001<sup>b</sup> |
| Missing N(%) | 126 (4.7%) | 2 (1.6%) |  |
| Cognitive Score |  |  |  |
| Mean ± SD | 78.3 ± 19.2 | 75.9 ± 17.7 | 0.262<sup>b</sup> |
| Missing N(%) | 1075 (40.1%) | 48 (38.4%) |  |
| ADOS Comparison Score |  |  |  |
| Mean ± SD | 7.1 ± 2.2 | 7.2 ± 2.4 | 0.482<sup>b</sup> |
| Missing N(%) | 485 (18.1) | 21 (16.8) |  |
| ADOS SA Relative |  |  |  |
| Mean ± SD | 7 ± 2.5 | 7.2 ± 2.6 | 0.440<sup>b</sup> |
| Missing N(%) | 479 (17.8) | 21 (16.8) |  |
| ADOS RRB Relative |  |  |  |
| Mean ± SD | 7.2 ± 1.9 | 7.5 ± 1.8 | 0.137<sup>b</sup> |
| Missing N(%) | 466 (17.3) | 20 (16) |  |
| DSM-5 Support Level Severity A (1)* N(%) | 438 (22.5%) | 20 (17.1%) | 0.002<sup>c</sup> |
| DSM-5 Support Level Severity A (2)* N(%) | 888 (45.6%) | 41 (35%) |  |
| DSM-5 Support Level Severity A (3)* N(%) | 621 (31.9%) | 56 (47.8%) |  |
| Missing N(%) | 733 (37.6%) | 8 (6.8%) |  |
| DSM-5 Support Level Severity B (1)* N(%) | 494 (25.3%) | 22 (18.8%) | 0.001<sup>c</sup> |
| DSM-5 Support Level Severity B (2)* N(%) | 1011 (51.8%) | 52 (44.4%) |  |
| DSM-5 Support Level Severity B (3)* N(%) | 444 (22.8%) | 43 (36.7%) |  |
| Missing N(%) | 731 (37.5%) | 8 (6.8%) |  |

<sup>a</sup>Chi-squared test; <sup>b</sup>T-test; <sup>c</sup>Cochran-Armitage test for trend; \*DSM severity level: 1- Requiring support, 2 - Requiring substantial support, 3 - Requiring very substantial support;

### ES analysis

ES analysis was performed using Illumina HiSeq sequencers and Illumina Nextera exome capture kit, following construction of BAM/CRAM files (human genome build 38) and variant calling format (vcf) files using the Genome Analysis Toolkit (GATK)^19^ or Illumina’s DRAGEN pipeline^20^. Then, we used our in-house bioinformatics pipeline *Autscore*^21^ for selecting only rare (< 1%), proband-specific (e.g. de-novo) and gene-disrupting variants. Finally, the clinical relevance for ASD of these candidate variant were evaluated by the ANCAN’s clinical genetic team according to the standard ACMG/AMP guidelines^22^. Consequently, clinically relevant genetic variants were found in 36 children.

### Developing a standardized ASD phenotype battery

Phenotypic data of participating children were obtained from two sources: 1) the ANCAN database that contains comprehensive sociodemographic and behavioral data on participating children; and 2) SUMC medical records, which encompass detailed clinical data on core ASD symptoms, and co-occurring medical conditions. We included only behavioral symptoms and comorbid conditions with a chronic or recurring nature and excluded acute conditions that occurred only once or were not directly related to an underlying medical condition, such as injuries and fractures. In total, 246 phenotypic variables were collected for the study participants (**Table S1**).

Next, we used the HPO nomenclature to assign each phenotype its unique HPO term and arrange these HPO terms into clusters according to the HPO hierarchy system using the *ontologySimilarity* package in R^23^. We manually reviewed and adjusted the clusters to ensure they reflect the biological relationships of the terms. Consequently, we excluded one HPO term that appeared only once in our data and another five that did not cluster with other phenotypes. Another 14 HPO terms were excluded because they did not associate with any gene in the HPO database. The remaining 228 HPO terms were grouped into a battery of 41 phenotype clusters (**Table S1**).

### Examining the association of the ASD phenotype battery with ASD genes

To evaluate the association between the selected ASD phenotype battery and established ASD-related genes, we quantified, for each gene, the number of battery phenotypes associated with it in the HPO database. We then compared the distribution of these counts between ASD-associated genes and non-ASD genes, as defined by the HPO and the SFARI Gene databases^24^. Statistical significance of the differences between groups was assessed using a two-sided t-test.

### Prioritization of candidate variants

To enable prioritization of ASD candidate variants using the phenotype battery, each participant was represented by an OPV, and each gene in the HPO database was represented by an EPV derived from its annotated HPO terms. Genotype–phenotype (EPV-OPV) similarity was then quantified for each participant using two complementary approaches:

1. We calculated the *SimGIC* score^25^, which is defined as the sum of the information content (IC) of the shared HPO terms (t) of the participant (P) and the gene (G) divided by the union of HPO terms – both computed from the ontology’s topological structure using the *descendants_IC* function in *ontologySimilarity* package (the frequency with which a term serves as an ancestor to other terms).

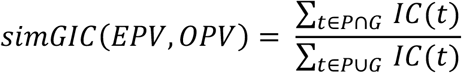
2. We employed *VarElect*, a commonly used prioritization tool that utilizes the GeneCards platform to associate mutated genes with relevant disease phenotypes with the phenotypes of our ASD phenotype battery.

Finally, we applied the *simGIC* approach to the candidate variants in the 89 children with unknown genetic causes. To enable practical use of the phenotypic battery and reduce the number of candidate variants requiring clinical evaluation, we defined an optimal *simGIC* threshold above which genes were considered high-priority. Threshold optimization was determined based on the *simGIC* results in the 36 genetically resolved children using the *Fβ* score, which is a weighted harmonic mean of precision and recall that enables explicit control over the relative importance of false positives and false negatives^26^.

## Results

A total of 125 children (76% males, 76.8% Jewish) were included in the study sample. Their basic sociodemographic and clinical characteristics in comparison to the other children in the ANCAN database are presented in **Table 1**. The study sample had a slightly higher percentage of Bedouin children (21.6% vs. 13.3%; p=0.025), was much younger at ASD diagnosis (37.5±17.6 vs. 52.9±37 months; p<0.001), and required more support according to the DSM-5 criteria (p=0.002 for DSM-5 A criteria and p=0.001 for DSM-5 B criteria; **Table1**). These differences may reflect greater motivation among parents of children with more severe symptoms who tend to be diagnosed earlier, to pursue early diagnosis and participate in genetic testing, potentially influenced by considerations for future family planning.

### Observed phenotypes among children with ASD

Overall, 228 distinct HPO terms, organized into 41 phenotype clusters, were identified in the study sample (**Table S1**). The largest cluster, “abnormality of the face” (HP:0000271), comprised 29 HPO terms, whereas ten clusters (e.g., “anxiety”; HP:0000739) each included a single term. **Figure 1A** illustrates the distribution of these 41 phenotype clusters across the 125 participants. The phenotype battery captured substantial inter-individual variability, with each participant exhibiting a unique phenotypic profile and a median of nine phenotypes per individual. **Figure 1B** compares phenotype prevalence between the study sample and HPO genes (defined as the proportion of HPO genes annotated to a given phenotype). Overall, 13 phenotypes (31.7%) were significantly more prevalent in the ASD sample than in the HPO gene set (p<0.05).

**Figure 1.**
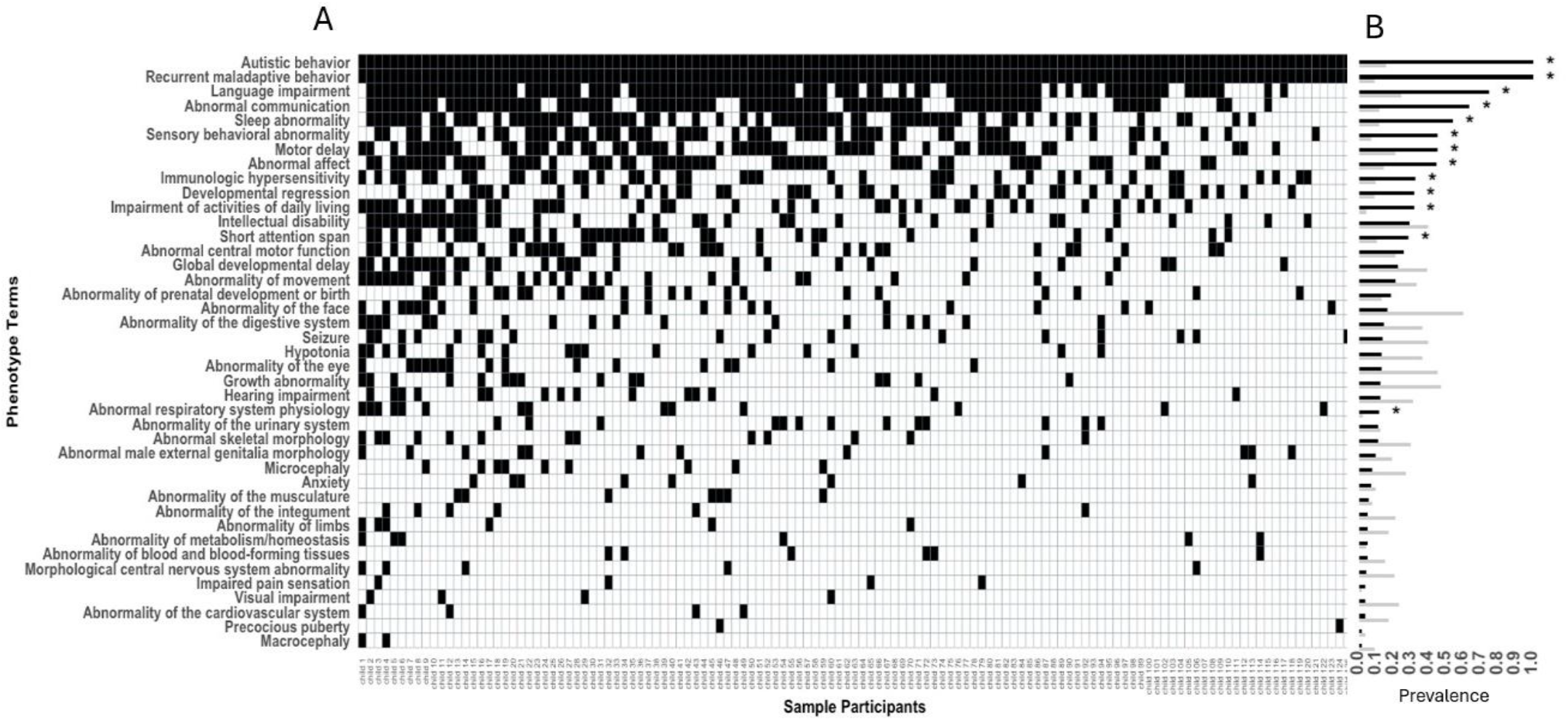
The ASD phenotype battery. **(A)** A matrix of the 41 HPO broad phenotypes (Y-axis) distributed across 125 participants (X-axis). Black cells indicate the presence of a specific phenotype for each participant. **(B)** The relative prevalence of the 41 phenotypes in the study sample (Black bars) and in the HPO database (Grey bars). * Indicates significant differences (p < 0.05) using chi-square test.

### Association of the ASD phenotype battery with known ASD genes

Figure 2. presents the distribution of the ASD battery’s phenotypes associated with ASD and non-ASD genes according to either the HPO or SFARI gene databases. In the HPO database (**Figure 2A**), ASD genes were associated with an average of 16.1 ± 5.7 phenotypes per gene compared to only 6.5 ± 5.4 phenotypes among non-ASD genes (p = 1.1^e-231^). Similarly, genes defined as “high confidence ASD genes” or “strong candidate or suggestive evidence” genes in the SFARI Gene database (**Figure 2B**) were associated with a significantly higher number of phenotypes than non-ASD genes (16.0 ± 6.7 and 10.4 ± 7.3 vs. 7.3 ± 6.0; p = 2.1^e-57^ and p = 1.9^e-14^ respectively). These large and highly significant differences reinforce the relevance of our phenotype battery to ASD genetics. Notably, several non-ASD genes were associated with relatively high number of phenotypes (**Figure 2**), thus highlighting them as potential novel ASD genes.

### Prioritization of candidate ASD variants

To assess the performance of our ASD phenotype battery in prioritizing ASD candidate variants, we applied both the *SimGIC* and *VarElect* approaches to the ES data of the 36 genetically resolved children in our sample. Of the 1,035,288 variants in the raw vcf data, 560 variants passed the *Autscore* filtering and considered as candidate variants (Mean = 15.6, Median = 13.5 per individual). **Figure 3** displays the ranking of these variants based on the similarity of their expected phenotypes to the observed phenotypes in each child using either *SimGIC* or *VarElect* approaches. Notably, both approaches yielded very similar results with *SimGIC* performing slightly better than *VarElect* by ranking the true clinically relevant variant as 1^st^ in 21 children (58%) compared to 19 (53%) children and in the top three variants in 32 children (89%) compared to 29 children (81%) respectively.

**Figure 2.**
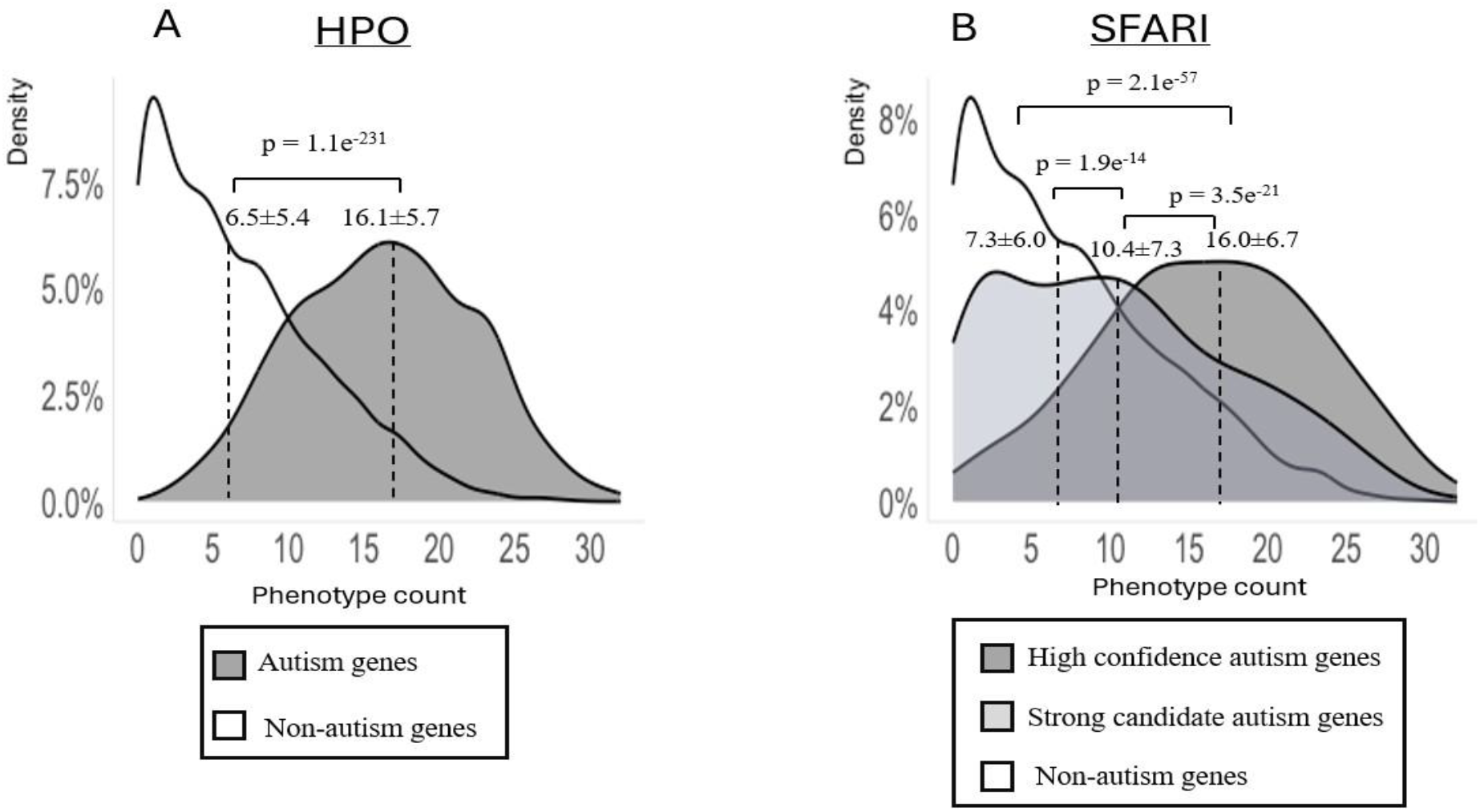
Comparison of phenotype counts between ASD and Non-ASD genes. **(A)** Density plots of the phenotype counts associated with ASD genes (n=780; Grey plot) and non-ASD genes (n= 4,348; Grey plot) according to the HPO database. **(B)** Density plots of the phenotype counts associated with “high confidence” ASD genes (dark Grey plot; n=268), “strong candidates” ASD genes (light Grey plot; n=372), and non-ASD genes (White plot; n=4,488), according to the SFARI Gene database.

**Figure 3.**
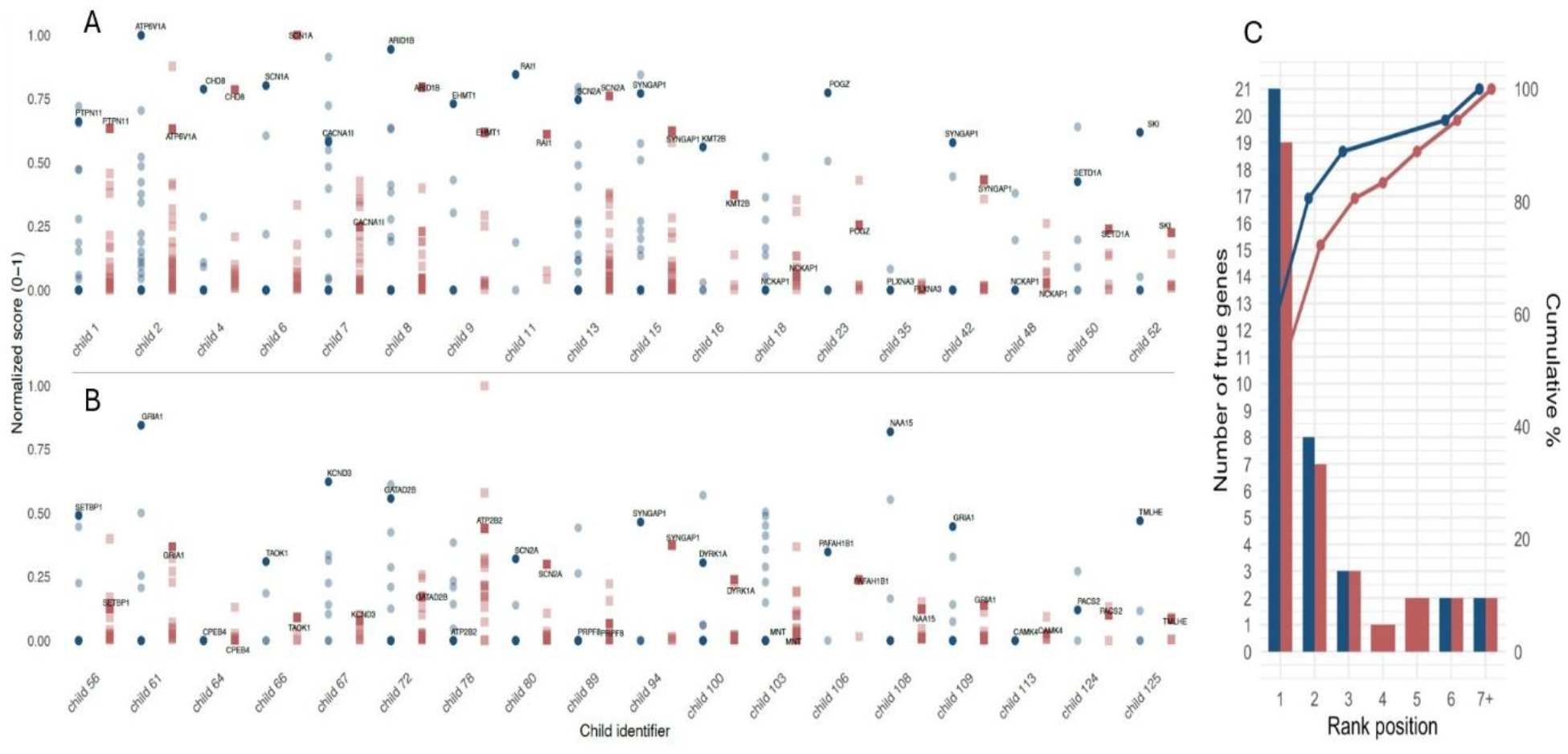
Prioritization of candidate variants across 36 children with known clinically relevant variants (resolved cases). **(A&B)** For each case, the normalized prioritization score (ranging from 0 to 1) of either *SimGIC* (Blue circles) or *VarElect* (Red squares) of candidte variants are plotted. indicate known with gene symbols indicate the clinically relevant variant in each solved case. **(C)** Bar and cumulative plot of the ranking position of clinically relevant variants in the 36 resolved cases by both *SimGIC* (Blue) or *VarElect* (Red). Overall, the clinically relevant variant was ranked 1^st^ in 21 (58%) and 19 (53%) of the cases and among the top-three variants in 32 (89%) and 29 (81%) of the cases using *SimGIC* and *VarElect* tools respectively.

We next applied the *SimGIC* approach to the 1,412 candidate variants in the ES data of the 89 genetically unsolved cases. The results are summarized in supplementary **Table S2**. Overall, 125 variants in 56 children exceeded the predefined *SimGIC* threshold (≥0.29), representing an 11.3-fold reduction in the number of variants requiring clinical evaluation. Subsequent review by our clinical genetics team identified six novel clinically relevant variants in five individuals, increasing the diagnostic yield in this sample by 45% (from 11 to 16 cases). Additional ten variants were classified as strong candidate variants by the team.

## Discussion

We present here a novel ASD phenotype battery comprised of 41 broad HPO phenotype categories. We show that these phenotypes are significantly enriched in known ASD genes, thus underscoring their association with ASD genetics. We further show that integration of this phenotype battery in the analysis of ES data may substantially improves genetic diagnosis of ASD by highlighting new clinically relevant variants undetected by standard approaches.

The broad range of 41 phenotypes captured by the battery highlights the substantial clinical heterogeneity of ASD beyond its core social-communication and behavioral features. The phenotypic variability was evident even among individuals carrying variants in the same gene (e.g., *SCN2A*), further underscoring the complexity of ASD.

Approximately one-third of the phenotypes included in the battery were enriched in the ASD sample relative to their frequency among HPO-annotated genes. These phenotypes primarily represented core ASD features and common neurodevelopmental and behavioral comorbidities, such as intellectual disability and sleep abnormalities. However, they also included immune and respiratory phenotypes, consistent with reports of increased rates of allergic and respiratory conditions among individuals with ASD^27–29^. The remaining two-thirds included several well-established ASD-associated comorbidities^30–34^, including features recently identified as predictors of a likely pathogenic or pathogenic genetic diagnosis in individuals with ASD^8^. The lack of significant enrichment for these phenotypes should not be interpreted as evidence of limited relevance to ASD, but may instead reflect their lower prevalence, limited statistical power, variability in clinical ascertainment, or differences in the completeness of HPO gene annotations.

The enrichment of phenotypes captured by our battery among established ASD genes in both the HPO and SFARI Gene databases, together with the graded pattern observed across SFARI evidence tiers, supports the biological and clinical relevance of our phenotypic framework. Notably, several genes not currently classified as ASD-related exhibited a relatively high burden of battery phenotypes, suggesting that they may represent underrecognized contributors to ASD susceptibility.

Using the phenotype battery to prioritize candidate variants with either *SimGIC* or *VarElect* proved highly effective, placing the clinically relevant variant among the top three candidates in 89% and 81% of genetically resolved children, respectively. This strong performance demonstrates the value of integrating detailed clinical phenotyping into the interpretation of ES data in ASD. Application of the SimGIC-based approach to genetically unresolved cases identified six additional clinically relevant variants in five individuals, increasing the number of genetically diagnosed cases by nearly 50%. These findings indicate that comprehensive, participant-specific phenotyping can improve both the efficiency and diagnostic yield of genomic interpretation, particularly for cases that remain unresolved following conventional variant filtering. Nevertheless, because the approach was developed and evaluated in the same relatively small cohort, its performance and the optimal similarity threshold require validation in independent and more diverse ASD populations.

From a clinical perspective, our findings highlight the potential utility of this ASD-specific phenotype battery as a front-line tool to inform genetic evaluation even prior to formal genetic testing. Systematic characterization of a child’s phenotypic profile using this battery may enable clinicians to generate a prioritized set of candidate genes based solely on observed clinical features, thereby guiding decision-making before genetic testing is initiated. By translating phenotypic data into a structured representation and leveraging established genotype–phenotype associations (e.g., through resources such as the HPO and SFARI Gene), clinicians could identify likely molecular etiologies and tailor subsequent diagnostic strategies, including selection of targeted gene panels, prioritization of variant interpretation, or early referral for comprehensive genomic testing. This approach may be particularly valuable in settings where access to sequencing is limited and could help streamline the diagnostic trajectory by focusing attention on the most plausible genetic contributors. Beyond improving efficiency, early phenotype-driven gene prioritization may facilitate anticipatory clinical management by highlighting gene-specific risks and comorbidities even before molecular confirmation. Collectively, these findings support the integration of structured phenotyping frameworks into ASD diagnostic workflows as a means to enhance precision medicine approaches from the earliest stages of clinical evaluation.

The results of this study should be interpreted under its limitations. First, the HPO database has an incomplete gene coverage and annotation which limits its ability to prioritize certain causal genes, particularly those lacking phenotype annotations or with insufficient representation of neurobehavioral features. Nevertheless, utilization of this battery is not restricted to the HPO and can be applied to other resources such as *VarElect* as demonstrated in this study. Second, variability and incompleteness in phenotypic data, arising from under-recognition of subtle traits (e.g., dysmorphic features, head circumference abnormalities, or specific seizure types), differences in physician documentation, and reliance on non-standardized clinical records or ICD-9 codes, may have affected phenotype capture and accuracy. In addition, the timing of clinical assessments may have contributed to misclassification, as some genotype-associated features emerge later in development and may not be evident at the time of evaluation. Thus, further improvement of the phenotype battery in these aspects may enhance the variant prioritization capacity of this approach.

## Conclusion

Our findings underscore the value of phenotype-driven frameworks for advancing the genetic diagnosis of ASD. By incorporating a broad range of clinical features beyond the core diagnostic domains, this approach can increase diagnostic yield and support the identification of novel candidate genes. More broadly, integrating comprehensive phenotypic data with genomic analyses may improve the interpretation of ASD-associated variation, refine etiologically meaningful subgroups, and ultimately inform more individualized approaches to clinical care.

## Data Availability

All data produced in the present study are available upon reasonable request to the authors

## Acknowledgements

We thank the families who participated in this study.

## Ethics Declaration

This study was approved by the ethics committee of SUMC (#039-21). Informed consent was obtained from all participants as required by the ethics committee.

## Author contributions

Conceptualization: N.L., I.M.; Data curation: N.L., M.D.; Formal analysis: N.L.; Investigation: N.L., M.D., M.I., D.Z., G.M., A.M.; Resources: G.M., A.M.; Funding acquisition, Project administration, and Supervision: I.M; Writing-original draft: N.L.; Writing-review & editing: I.M.

